# RESIDENTS’ PERCEPTION ON CENTRAL STORAGE OF SOLID WASTES IN ILORIN METROPOLIS, KWARA STATE. NIGERIA

**DOI:** 10.1101/2022.02.14.22270939

**Authors:** Yusuf Olanrewaju Raufu, Adewoye Solomon Olayinka, Sawyerr Henry Olawale

## Abstract

**Background:** Solid waste management in Nigeria has been a major concern to stakeholders due to inefficient collection and unsafe disposal. In Ilorin, the capital of Kwara state, the problem of wastes is becoming alarming because the more these wastes are evacuated the more they are generated on a daily basis. Thus, the issue of central storage of solid waste situation in Ilorin to say the least, is becoming quite distressing.

**Objectives:** This study addressed issues relating to waste management and perception of residence to central waste storage in Ilorin metropolis.

**Methods:** A cross-sectional study was conducted among the residents in the three local government that constitute Ilorin metropolis, residents living close to central wastes storages in Ilorin metropolis are purposively chosen and the registered wastes contractors with the Kwara State Environmental Protection Agency (KWASEPA). In accomplishing these objectives, both primary and secondary data sources were used. The primary data were collected via questionnaires and field observations. Secondary data were extracted from different published and unpublished materials. 120 households located on the main road and close to central wastes storage bins and 30 wastes contractors are administered with questionnaires. The data were analyzed using statistical package for social science (SPSS version 23) for descriptive and inferential at 5% level of significance.

**Results:** The operational capacities of wastes contractors showed a low capacity as 66.7% (20) operate with single rickety truck. Dumpsites accessibility and insufficiency is an impediment as the dumpsite was found to be too far and largely inaccessible. Condition of wastes spillage from the central storage and storage on the road meridians are draw backs to effective wastes management and majority of respondents 66.7% are unhappy.

**Conclusions:** It was discovered that central wastes storage system in Ilorin metropolis was not effective with constant incidence of wastes spillages and storage on road meridians. Therefore, capacity and competence should be considered in wastes management contract award, provision of more central wastes storage, establishing wastes transfer station, provision of adequate dumpsites at different local government, public enlightenment, regular trainings for wastes consultant, provision of loans for wastes consultants for acquisition of trucks and proper monitoring by regulatory agency will ensure a clean and sanitary environment in the metropolis.

## Introduction

Wastes are all things we consider unfit, unwanted and discarded due to economic reasons or ignorance of alternative technologies to re-use them [1, 2]. According to Adegoke [3], wastes are substances and materials that are disposed of or are required to be disposed of according to the provision of National Law. Wastes are abandoned as useless, yet if incorrectly processed, stored, transported, or dumped, they have the potential to cause death, sickness, or harm to humans or ruin the environment [4]. As a result, garbage is a byproduct of life, and its creation is worldwide. Individual, familial, municipal, industrial, and developmental activities all generate waste [5, 6, 7, 8, 9, 10, 11, 12]. Because garbage is created on a regular basis, it is also thrown away indiscriminately in certain locations, with complete disregard for the environment or immediate surroundings, such as land, sewers, or water bodies [13, 14, 15, 16, 17, 18, 19, 20, 21, 22]. Organic components are naturally destroyed and returned to the life cycle. The inorganic or non-biodegradable components, on the other hand, it remains as residues for days or weeks depending on their chemical composition, and some of them become hazardous [23, 24, 25, 26, 27, 28, 29, 30, 31]. The problem is exacerbated by the fact that current science and technology contribute to the release of thousands of new consumer items onto the market via discoveries and inventions. As a result, we have problems in protecting the planet’s delicate ecosystems [9, 10, 32]. Waste control and management are treated seriously in modern cultures. Waste is a severe challenge in a developing country like Nigeria due to a lack of awareness of its nature and the skills required to handle it properly. While industrialized nations are expanding their understanding and conscience about minimizing, reusing, and recycling garbage, certain countries, such as Australia and New Zealand, are heading toward zero waste to landfill. With anti-climate change slogans and efforts from concerned environmentalists and civic societies, no-burn technologies are becoming the norm [33, 34]. In Nigeria, the typical inhabitant generates 0.43kg of solid garbage every day. Organic waste accounts for 60 to 80 percent of total garbage, with plastics/nylon and scrap metal being the most recyclable elements [35, 36]. Waste management has been one of the major urban management challenges in Sub Saharan African countries and Nigeria is no exception. The challenges of management of solid waste which had turned some major cities into ugly sights to behold and resulted in the poor aesthetic appeal of those cities are the result of poor mastering of concepts, approaches and techniques [37]. Imam *et al*. [38] reported that piles of waste are dumped by the roadside and other open spaces, posing an environmental risk. Ogbonna *et al*. [39] observed that cities are divided into sections for the local contractors. However, inefficiency still thrives due to the lack of coordination by the Government and the lack of expertise on waste management issues by the environmental agencies.

## Methods

### Study Area

The study area is the city of Ilorin which is the largest urban center in Kwara State. It is also the capital of Kwara State. Ilorin city occupies an area of 89km^2^ and lies between the latitude 8.1333 and longitude 4.8833 along equator (see figure 1 below). Ilorin metropolis is composed of three local government areas; Ilorin West, Ilorin East, and Ilorin South. Ilorin city is the commercial and administrative center of the state [40, 41, 42]. The population of Ilorin according to 2006 census is 777,667 and this study covers all the three Local Government Councils.

**Figure 1:**
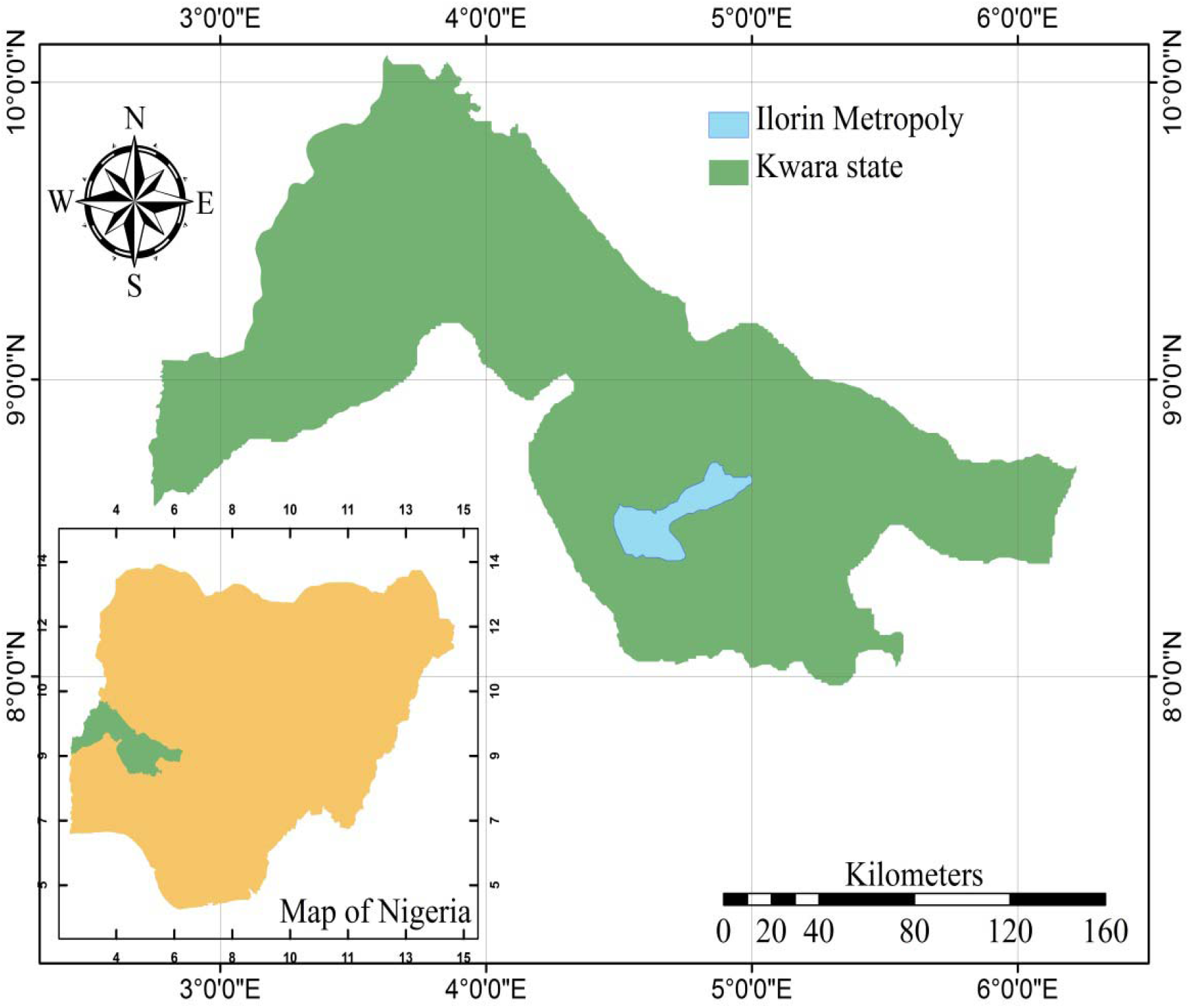
Map of Kwara State showing the study area. Adapted from Yusuf *et al*., [43]. [doi: https://doi.org/10.1101/2022.01.26.22269849.]

### Study Population

The study population are residents living close to central wastes storage and wastes management contractors working in Ilorin metropolis which is the capital city of Kwara State, Nigeria.

### Sampling Techniques

The sampling technique employed in this study is the purposive sampling technique. Only households identified as located in areas where there are central wastes storages bins and waste management contractors registered with government are selected for the study.

### Data Instrument

A semi-structured, interviewer administered questionnaires were used to elicit information on the socio-demographics and perception about central wastes storage in the city. The instrument was pre-tested using 12 households at Tanke area from residents along central wastes storage places. Each question was translated into the local languages for those that could not read English, to help the respondents to give true and accurate answers.

### Data Analysis

Data was analyzed using SPSS 23, using descriptive statistic such as mean, frequencies, charts and graphs.

### Ethical Approval

Ethical clearance for this study was obtained from the Kwara State Ministry of Health Ethical Review Committee. Informed consent of participants were sought before taking part in the study.

## Results and Discussion

### Socio-Demographic Characteristics of respondents

Here we present the results of the distribution of residents based on their age, marital status, educational level and tribal status respectively.

The Table 1 above shows results from wastes scavengers’ socio-demographic data. Also, figure 2 is the map of distribution of central wastes bins and number of bins within Ilorin metropolis, this covered three local government areas that made up the metropolis namely Ilorin west, Ilorin south and Ilorin east. The least number of central wastes bin at a place is 1 bin which occur in seven locations while the highest number of bins are 6 central wastes bins which are just two areas. Table 2 above show distribution of residents’ types and perception on effectiveness of central wastes storage and disposal. The residents who took part in the study cut across almost all strata of the society from residential, business premises, hospitality industries, and those that produce special wastes like hospitals, pharmacies, and diagnostic centres classified as others. The distribution is such that households constitute that majority with (65) representatives which is equivalent to 54.2% of the total respondents’ population size. Business premises and offices are next in terms of population with (27) representatives, constituting 22.5%. The next premise is termed others and this includes all areas that generate special waste and those under this category are the hospital, clinics, pharmacies and diagnostic centres. Finally, others comprise (17) representatives who make up 14.2% of the total population. On the closeness of central storage bins to premises, it was revealed that majority of the central storage bins are not located close to premises as 86 of respondents which was 71.77% of respondents stated that the central wastes storage bins are not located too close to their premises while 34 respondents which was 28.3% revealed that the central wastes storage bins are located too close to their residence. On adequacy of the central wastes’ storage bins, majority of residents believed the wastes storage bins are not adequate in quantity as 79 of the respondents which was 65.8% of the respondents while 23 which 18.7% are not sure and those that stated that the central wastes storage bins are adequate are just 18 which was 15% of the respondents. Figure 3 is a chart showing occurrence of wastes spillage from central wastes storage bins across the metropolis. Majority of the residents indicated occurrence of wastes spillage in the central waste’s storage point near them. The total number of 80 households, which is 66.7% of the respondents, revealed wastes spillages in the central storage in their areas. Those who are not sure of wastes spillages in their areas are 24, constituting 20% of the respondents. The number of respondents said there are no wastes spillages from central wastes storage in their areas are 16 which was 13.3% of the respondents. Figure 4 shows the environmental health concerns of central wastes storage each cluster is representing unique environmental health issues observable by the residents in the central wastes storage system in the metropolis. The first environmental health issue is flies’ infestation; questions are asked from the residents on flies’ infestation occurrences in their area. The results showed that the majority of the residents, 65% of them, which is equivalent to 78 households out of 120 households, have concerns concerning flies’ infestation because of fear of disease transmission. On the other hand, some of the households are indifferent to the issue of flies’ infestation and indicated that they are not sure if those flies pose any danger to them or their neighbourhoods. The respondents in this category constitute 20% of the respondents, which is equivalent to 24 households out of the total 120 respondents. The least categories believed that flies infestation could not constitute any health risks to them or their neighbourhoods. These groups comprise 15% of the total respondents, equivalent to 18 households. Also, among the observable issues of environmental health concern in the central wastes storage system is terrible odour from the central wastes storage areas that affect residents and passersby. Odour nuisance is another environmental health issue examined from questions asked from respondents. The information received showed that 60% of 72 households responded that odour nuisance constitutes a threat to the health of the residents and passersby. The remaining household that are not sure if the odour nuisance constitutes environmental health risk to the residents constitute 20% of the households which is equivalent to 24 respondents and this is similar to the number that opined that odour nuisance has no negative implication to the households and passerby which was also 20% of the respondents which amount to 24 households. Improper waste storage has been known to depreciate the aesthetic value of the environment and constitute a visual nuisance to the residents and visitors. Residents were asked of their perception of the central wastes storage in terms of depreciation of aesthetic value of the environment. The responses of the households to the questions as presented in figure 4 above showing that majority of the people, 43.4%, do not feel that central wastes storage constitute a visual nuisance, inform depreciation of the aesthetic values of the environment. On the other hand, those that feel the central waste storage devalue the city’s aesthetic appeal are 33.3% of the population, which amount to 40 households out of the total 120 households. The remaining 23.3% are not sure if the central wastes storage depreciates the aesthetic values of the environment or not. Figure 5 above present wastes storage on road meridian in Ilorin metropolis. The occurrence of wastes storage on the road divider or road meridian is becoming a new normal in the metropolis. Therefore, the researcher made enquiries from the residents to have their opinion on the practice. The result from the households is presented in figure 5 above. However, most residents opined that such practice is terrible and that can be seen from their responses because 87 out of 120 households constituting 72.5% of the population thought the practice was terrible. The number of residents who see nothing wrong in storing wastes on the road meridian are 19 out of 120 households, constituting 15.8% of the total population. Thus, those who cannot say if the practice is good or bad are classed as am not sure group; they are the minority among the respondents with 14 out of the 120 households, equivalent to 11.7% of the responding households. Figure 6 shows Safety concerns of central wastes storage in the metropolis. Central wastes storages are usually known to constitute some traffic issues, especially by way of wastes overflow that usually obstruct traffic flow; the researcher asked some pertinent safety questions from the respondents. The answers to some safety questions are presented in the chart below. According to the respondents, fire outbreak incidence is a very rare occurrence with 13.3% occurrence. The majority of households stated that they have not experienced fire outbreaks, while 65% of households said they have not seen or heard of any fire outbreak. The third group of respondents are households who are unsure of seeing or hearing of any fire incidents arising from central waste storage. Furthermore, visibility issues arising from central wastes storage across the streets in the metropolis has been of safety concern. Therefore, the residents across the metropolis were asked about visibility problems. On visibility issues, most households close to central wastes storage agreed that there are visibility issues with central wastes storage. The respondents who agreed that there is a problem of poor visibility due to central wastes storage are 51.7% of the total population while those who are not sure if central wastes storage constitute visibility issue are next with 25.8% of the population. Finally, the last group opined that central wastes storage did not constitute a visibility problem at any point in time. Those that stated that central wastes storage does not constitute a visibility problem are 22.5% of the total household involved in the study. Another safety concern raised in the questionnaire is traffic congestion problems. This is usually experienced due to overflow or spillage of wastes from the central wastes storage and during waste evacuation along the major streets in the metropolis. The residents’ perception of traffic congestion was revealed as presented in the chart above on the third stack of bars. Generally, most residents, constituting 69.2%, consider central wastes storage as no major cause of traffic problems in their areas. On the other hand, those who believe that central wastes storage is a major cause of traffic problems in their areas are just 15.8% and those that are not sure if central wastes storage constitutes traffic nuisance are a little less, which is 15% of the total respondents. The last safety issue examined was the occurrence of accidents due to the position of the central wastes’ storage. The respondents, mostly 56.7%, stated they had not seen or heard of any accident resulting from the location of the central wastes storage close to their residence. On the other hand, those that stated that they have experienced or witnessed the occurrence of accidents resulting from the position of the central wastes storage are % of the households and the remaining 20% of the population are not sure if they have heard or experienced the occurrence of accidents as a result of the position of the central wastes storage in their areas.

**Table 1:**
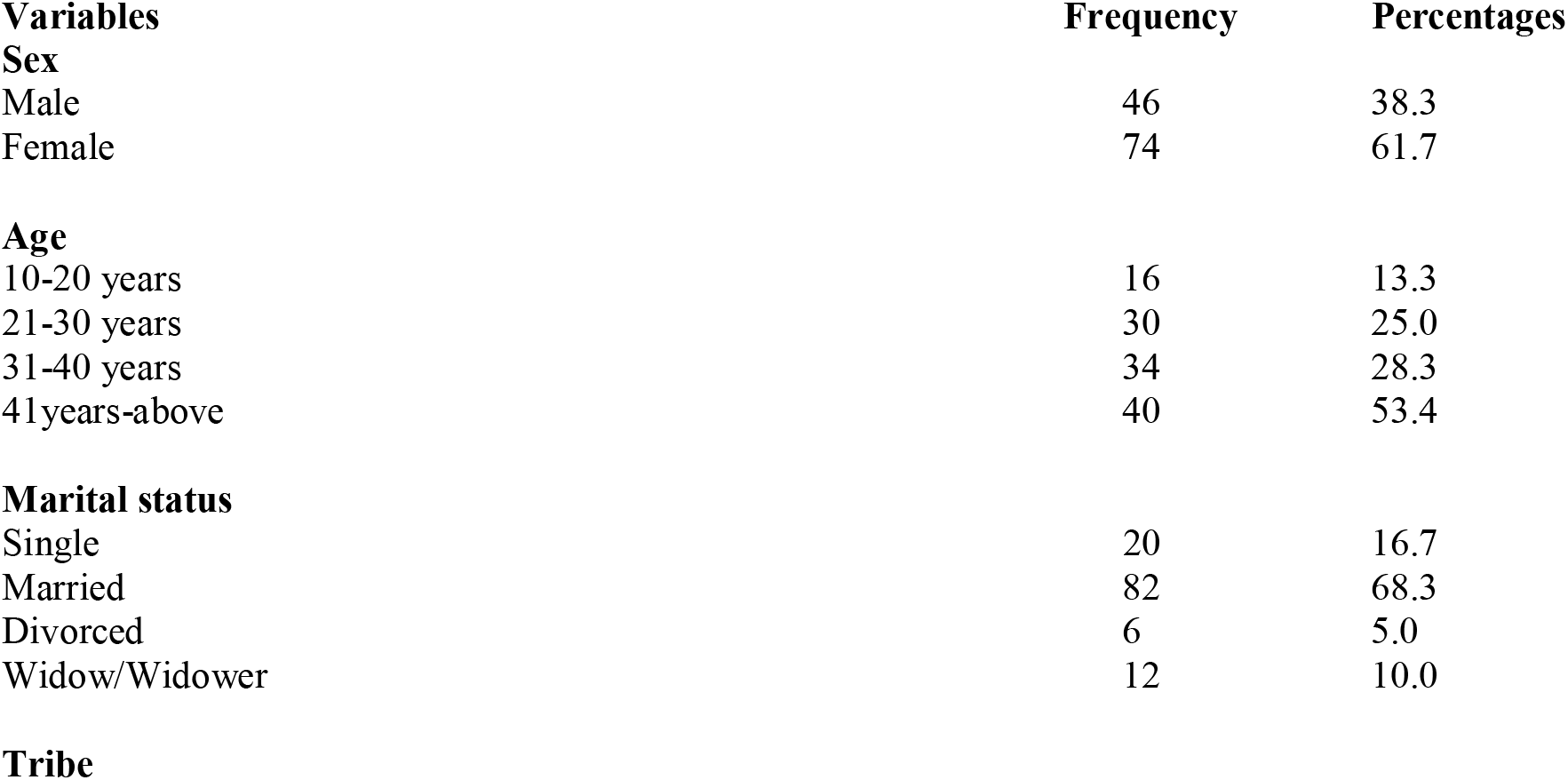
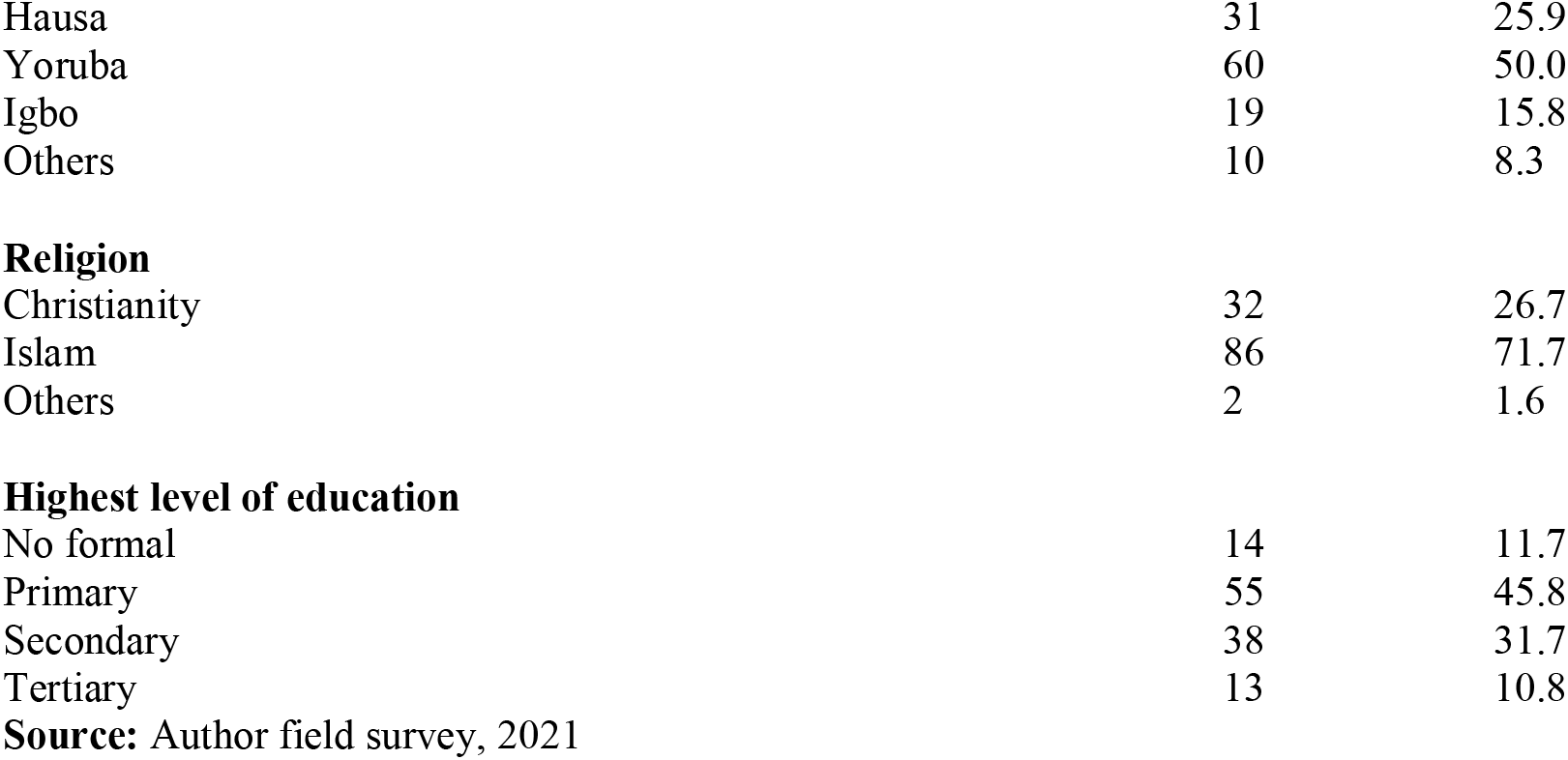
Demographic Characteristics of the Respondents

| **Variables** | **Frequency** | **Percentages** |
| --- | --- | --- |
| **Sex** |  |  |
| Male | 46 | 38.3 |
| Female | 74 | 61.7 |
| **Age** |  |  |
| 10-20 years | 16 | 13.3 |
| 21-30 years | 30 | 25.0 |
| 31-40 years | 34 | 28.3 |
| 41years-above | 40 | 53.4 |
| **Marital status** |  |  |
| Single | 20 | 16.7 |
| Married | 82 | 68.3 |
| Divorced | 6 | 5.0 |
| Widow/Widower | 12 | 10.0 |
| **Tribe** |  |  |
| Hausa | 31 | 25.9 |
| Yoruba | 60 | 50.0 |
| Igbo | 19 | 15.8 |
| Others | 10 | 8.3 |
| Religion |  |  |
| Christianity | 32 | 26.7 |
| Islam | 86 | 71.7 |
| Others | 2 | 1.6 |
| Highest level of education |  |  |
| No formal | 14 | 11.7 |
| Primary | 55 | 45.8 |
| Secondary | 38 | 31.7 |
| Tertiary | 13 | 10.8 |

**Source:** Author field survey, 2021

**Table 2:** Effectiveness of central wastes storage and Disposal

| **Premises Type** | **Frequency** | **Percentage** |
| --- | --- | --- |
| Household | 65 | 54.2 |
| Hotel/Eatry | 11 | 9.2 |
| Business Premise/Offices | 27 | 22.5 |
| Others | 17 | 14.2 |
| **Total** | **120** | **100** |
| **Central Storage Too Close** | **Frequency** | **Percentage** |
| Yes | 34 | 28.3 |
| No | 86 | 71.7 |
| **Total** | **120** | **100** |
| **Storage Adequate** | **Frequency** | **Percentage** |
| Yes | 18 | 15 |
| No | 79 | 65.8 |
| I Am Not Sure | 23 | 18.7 |
| **Total** | **120** | **100** |

**Figure 2:**
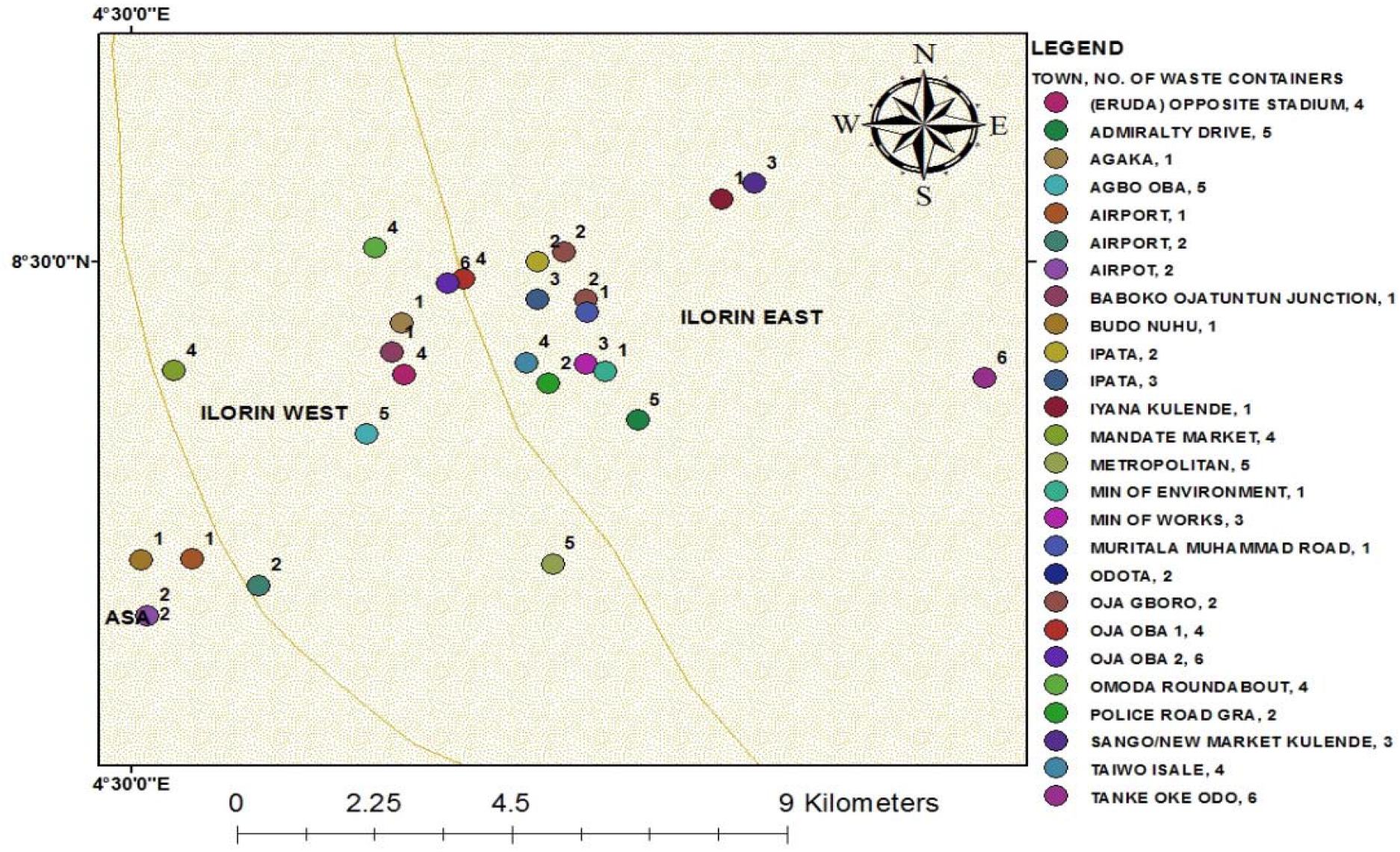
Map of distribution of central wastes bins and number of bins within Ilorin metropolis

**Figure 3:**
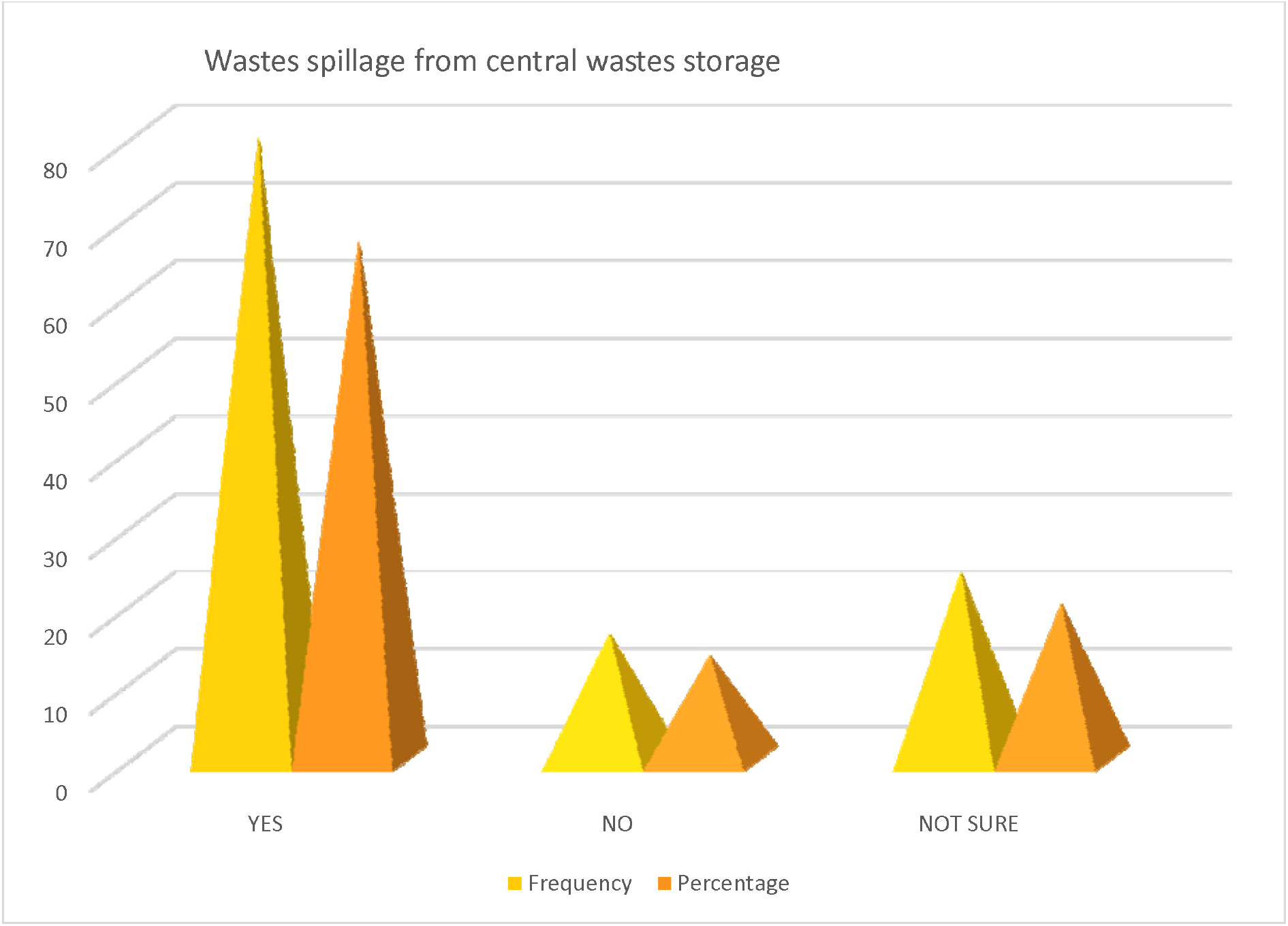
Wastes spillage from central wastes storage

**Figure 4:**
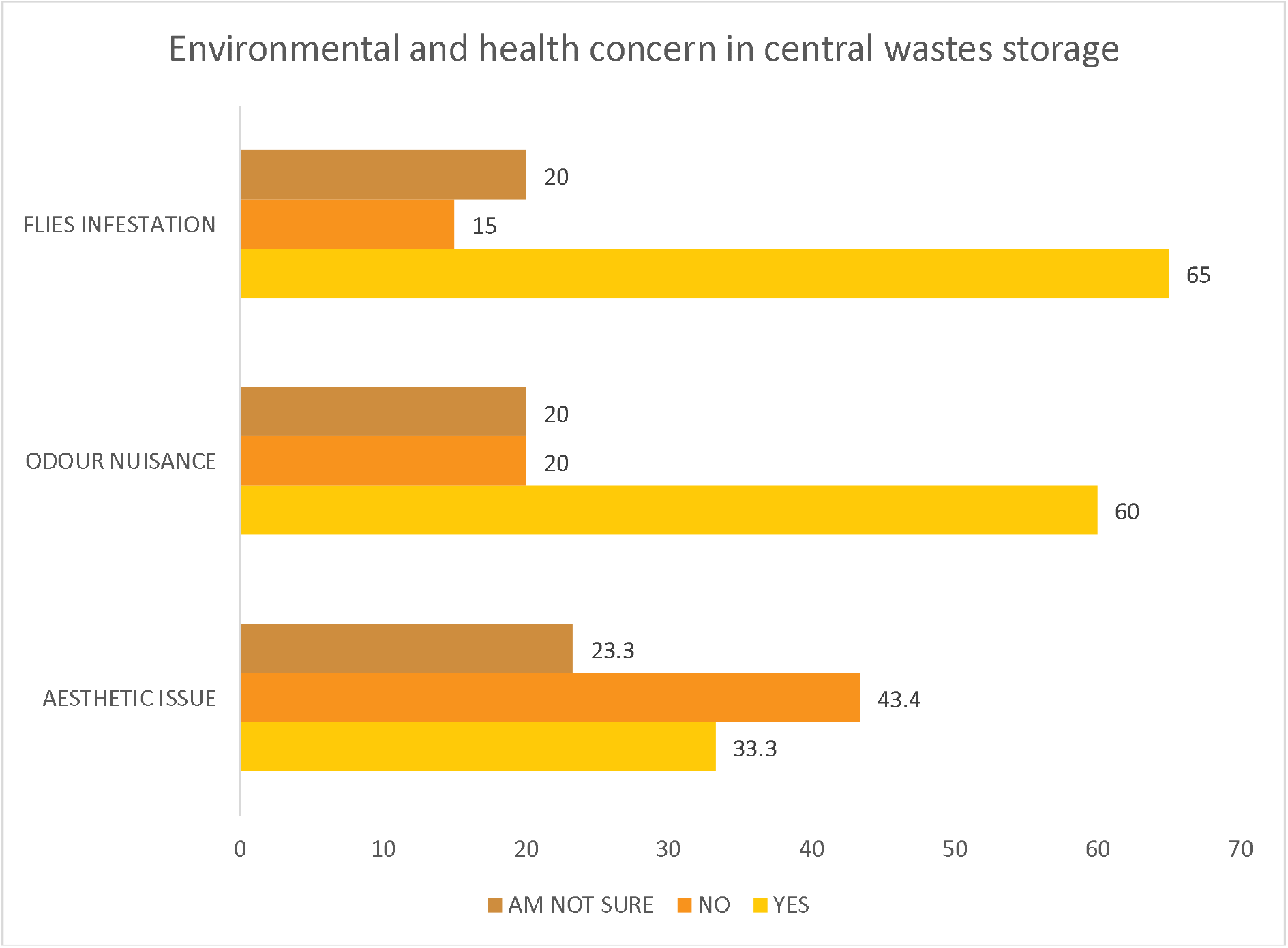
Environmental health concerns of central wastes storage

**Figure 5:**
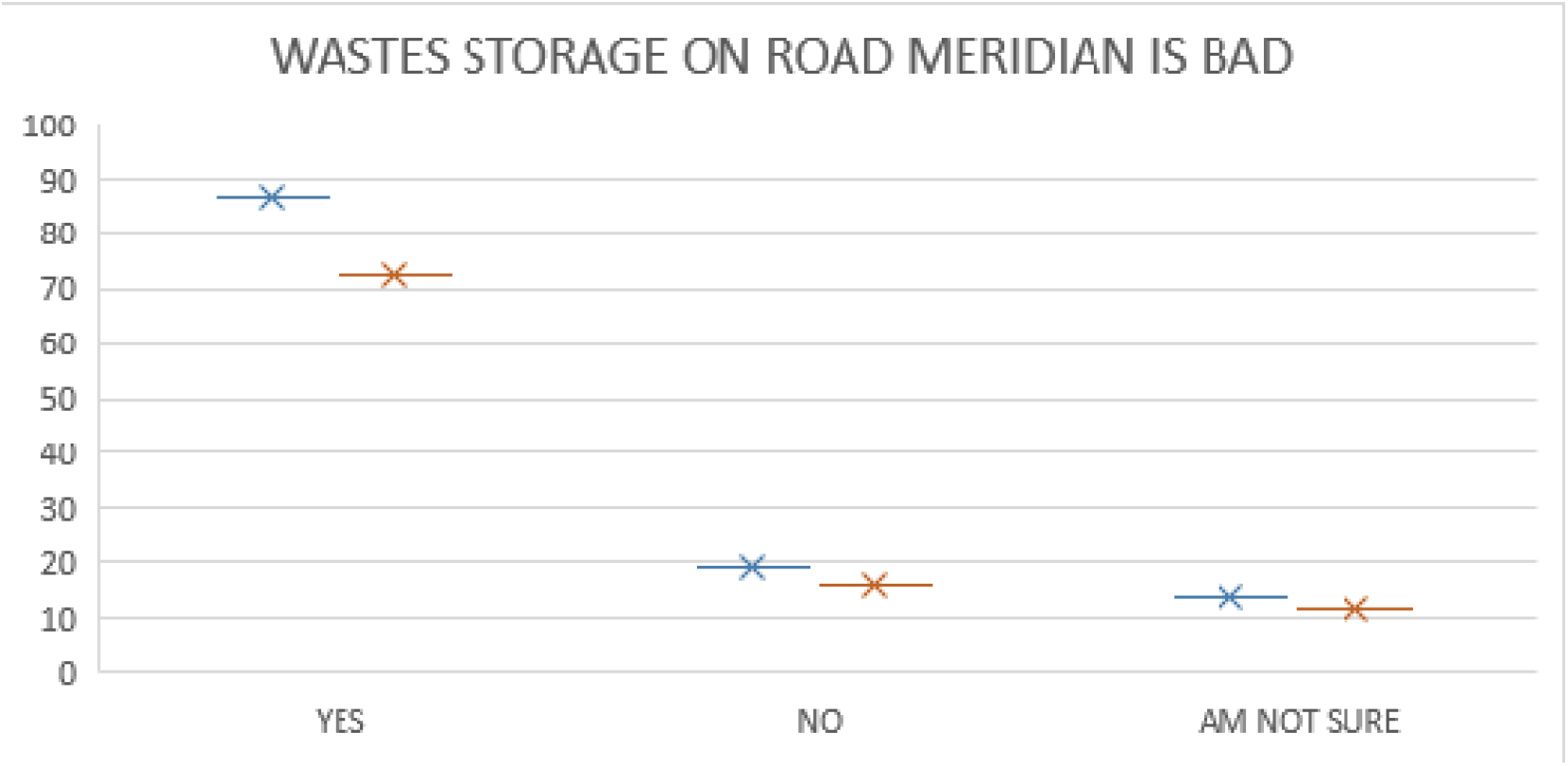
Wastes storage on road meridian in Ilorin metropolis

**Figure 6:**
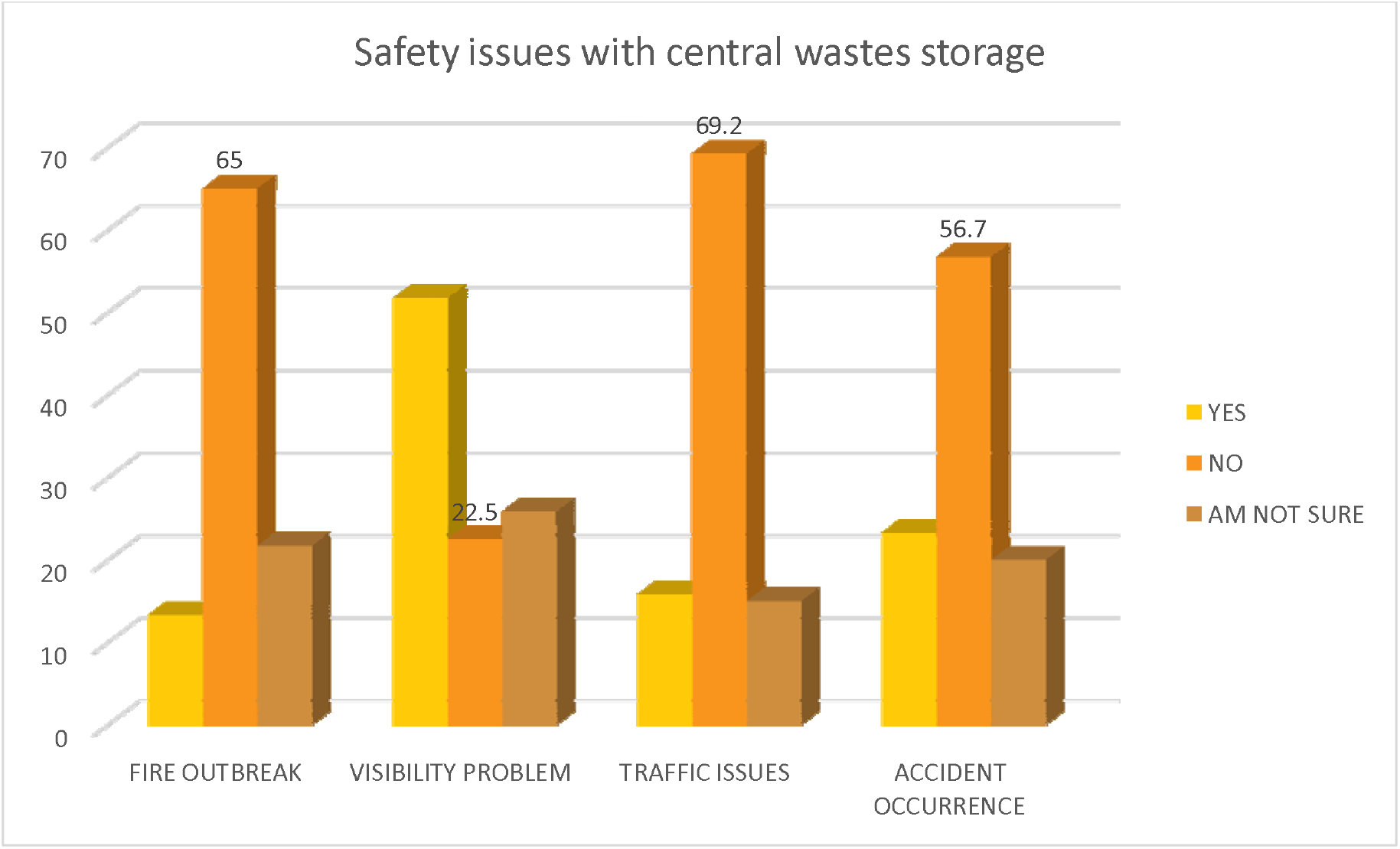
Safety concerns of central wastes storage

Summarily, it was found that central wastes storage distributions were insufficient and majorly found in high-brow areas and the highest number of wastes bins found in a location were 6 and the least being 1 and other places don’t have at all. This is in agreement with the findings of Batool [44] and Joseph [45] that stated that unavailability or insufficiency of dumpsters and poor transportation is a main drawback for efficient wastes management. Lack of sufficient waste collecting points was found as barrier by Nachalida *et al*. [46]. Also, wastes spillage from the central storage is one of the draw backs of the system and majority of the residents 66.7% are unhappy with it while only 13.3% of respondents don’t see spillage as a problem. This agreed with Kaoje *et al*. [47] on wastes storage situation in Sokoto state, 94.1% of resident are worried how solid waste litters the metropolis. On the storage of wastes on the road meridian and by the road sides, majority of respondents 72.5% opined that it is a bad practice. Fagariba and Song [48], also concluded that inadequate access to these waste bins as one of the main barriers. Issues relating to environmental health and safety concern of residents show majority of residents close to central wastes storage are concerned about flies’ infestation with 63% respondents and follow by odour nuisance while aesthetic is the least of their concern. This was also identified by Steven [49], who associated central wastes storage with flies’ nuisance and transmission of diseases and odours.

## Conclusion

In recent years, the topic of environmental management and protection has taken on a worldwide scale. It has become more crucial that man’s physical actions and activities favourably meet with necessary environmental norms and laws for the general well-being of people and society. To that purpose, different laws have been adopted at various times and are typically aimed at assuring sufficient management and preservation of the physical habitat, provided that good environmental management is legislatively driven. Thus, the public perception of solid waste management methods in Ilorin metropolis was researched in relation to central wastes storage in the metropolis, and it was discovered that citizens felt the existing wastes management practice could not handle the city’s wastes problem. Therefore, adequate provision of central wastes storage to prevent traffic obstruction, spillages, accidents and devaluation of aesthetic of the environment is recommended. Also, provision of adequate dumpsites at each local government should be considered and proper management of the dumpsite must be ensured. Establishment of transfer stations to ease the travel time of wastes companies. Material Recovery facility should be established to recover recyclables from the waste and boost the economy through wastes to wealth.

## Data Availability

All data produced in the present work are contained in the manuscript

## Ethics approval

Ethical approval was granted from the Kwara State Ministry of Health Ethical Review Committee. Informed consent was obtained from each respondent.

## Residents’ consent

Obtained

## Data Availability

Data used to support the findings of this study are included within the article.

## Acknowledgements

Authors are thankful to all participant who participated in this research study and also Sanitarian Raimi Morufu Olalekan for editing the article. This study was a part of a PhD Thesis of Yusuf Olanrewaju Raufu, (Department of Environmental Health Science, Kwara State University, Malete).

## References

1. Okebukola, PAO (2001): Our Environment, Our Destiny. A paper delivered at the distinguished lecture series of Adeniran Ogunsanya College of Education cited in STAN Environmental Education Series, No. 5. P 1.

2. Suleiman RM, Raimi MO and Sawyerr HO (2019) A Deep Dive into the Review of National Environmental Standards and Regulations Enforcement Agency (NESREA) Act. International Research Journal of Applied Sciences. pISSN: 2663-5577, eISSN: 2663-5585. DOI No. Irjas.2019.123.123. www.scirange.com. https://scirange.com/abstract/irjas.2019.108.125.

3. Tchobanoglous, G., Theisen, H. and Vigil, SA. (1993) Integrated Solid Waste Management: Engineering Principle and Management Issue. McGraw Hill Inc., New York.

4. Adegoke, OS. (1990): Waste Management within the Context of Sustainable Development. Department of Geology, Obafemi Awolowo University, Ile-Ife.

5. Raimi MO, Suleiman RM, Odipe OE, Salami JT, Oshatunberu M, et al (2019). Women Role in Environmental Conservation and Development in Nigeria. Ecology & Conservation Science; 1(2): DOI: 10.19080/ECOA.2019.01.555558. Volume 1 Issue 2 - July 2019. https://juniperpublishers.com/ecoa/pdf/ECOA.MS.ID.555558.pdf

6. Ebuete AW, Raimi MO, Ebuete IY & Oshatunberu M (2019) Renewable Energy Sources for the Present and Future: An Alternative Power Supply for Nigeria. Energy and Earth Science. Vol. 2, No. 2, 2019. URL: http://dx.doi.org/10.22158/ees.v2n2p18. http://www.scholink.org/ojs/index.php/ees/article/view/2124.

7. Ajayi FA, Raimi MO, Steve-Awogbami OC, Adeniji AO, Adebayo PA (2020) Policy Responses to Addressing the Issues of Environmental Health Impacts of Charcoal Factory in Nigeria: Necessity Today; Essentiality Tomorrow. Communication, Society and Media. Vol 3, No 3. DOI: https://doi.org/10.22158/csm.v3n3p1. http://www.scholink.org/ojs/index.php/csm/article/view/2940.

8. Okoyen E, Raimi MO, Omidiji AO, Ebuete AW (2020). Governing the Environmental Impact of Dredging: Consequences for Marine Biodiversity in the Niger Delta Region of Nigeria. Insights Mining Science and technology 2020; 2(3): 555586. DOI: 10.19080/IMST.2020.02.555586. https://juniperpublishers.com/imst/pdf/IMST.MS.ID.555586.pdf.

9. Olalekan MR, Abiola I, Ogah A, Dodeye EO (2021) Exploring How Human Activities Disturb the Balance of Biogeochemical Cycles: Evidence from the Carbon, Nitrogen and Hydrologic Cycles. Research on World Agricultural Economy. Volume 02, Issue 03. DOI: http://dx.doi.org/10.36956/rwae.v2i3.426. http://ojs.nassg.org/index.php/rwae.

10. Raimi MO, Abiola I, Ogah A, Dodeye EO and Aziba-anyam GR (2021) Exploring How Human Activities Disturb the Balance of Biogeochemical Cycles: Evidence from the Carbon, Nitrogen and Hydrologic Cycles. IntechOpen. DOI: http://dx.doi.org/10.5772/intechopen.98533. https://www.intechopen.com/online-first/77696. ISBN 978-1-83969-144-7. Published: December 1st 2021; ISBN: 978-1-83969-144-7; Print ISBN: 978-1-83969-143-0; eBook (PDF) ISBN: 978-1-83969-145-4. Copyright year: 2021.

11. Morufu OR, Aziba-anyam GR and Teddy CA (2021) ‘Silent Pandemic’: Evidence-Based Environmental and Public Health Practices to Respond to the Covid-19 Crisis. IntechOpen. DOI: http://dx.doi.org/10.5772/intechopen.100204. ISBN 978-1-83969-144-7. https://www.intechopen.com/online-first/silent-pandemic-evidence-based-environmental-and-public-health-practices-to-respond-to-the-covid-19-Published: December 1st 2021; ISBN: 978-1-83969-144-7; Print ISBN: 978-1-83969-143-0; eBook (PDF) ISBN: 978-1-83969-145-4. Copyright year: 2021.

12. Raimi, MO., Mcfubara, KG., Abisoye, OS., Ifeanyichukwu Ezekwe, C., Henry SO., & Raimi, GA (2021) Responding to the call through Translating Science into Impact: Building an Evidence-Based Approaches to Effectively Curb Public Health Emergencies [COVID-19 Crisis]. Global Journal of Epidemiology and Infectious Disease, 1(1). DOI: 10.31586/gjeid.2021.010102. Retrieved from https://www.scipublications.com/journal/index.php/gjeid/article/view/72.

13. Raimi MO and Sabinus CE (2017) An Assessment of Trace Elements in Surface and Ground Water Quality in the Ebocha-Obrikom Oil and Gas Producing Area of Rivers State, Nigeria. International Journal for Scientific and Engineering Research (Ijser): Volume 8, Issue 6, July Edition. ISSN:2229-5518.

14. Olalekan, RM., Omidiji, AO., Nimisngha, D., Odipe, OE. and Olalekan, AS. (2018). Health Risk Assessment on Heavy Metals Ingestion through Groundwater Drinking Pathway for Residents in an Oil and Gas Producing Area of Rivers State, Nigeria. Open Journal of Yangtze Gas and Oil, 3, 191–206. https://doi.org/10.4236/ojogas.2018.33017

15. Henry OS, Morufu OR, Adedotun TA & Oluwaseun EO (2019) Measures of Harm from Heavy Metal Pollution in Battery Technicians’ Workshop within Ilorin Metropolis, Kwara State, Nigeria. Scholink Communication, Society and Media ISSN 2576-5388 (Print) ISSN 2576-5396 (Online) Vol. 2, No. 2, 2019 www.scholink.org/ojs/index.php/csm. DOI: https://doi.org/10.22158/csm.v2n2p73.

16. Olalekan RM, Adedoyin OO, Ayibatonbira A, et al (2019). “Digging deeper” evidence on water crisis and its solution in Nigeria for Bayelsa state: a study of current scenario. International Journal of Hydrology. 2019;3(4):244 257. DOI: 10.15406/ijh.2019.03.00187.

17. Olalekan RM, Dodeye EO, Efegbere HA, Odipe OE. Deinkuro NS, Babatunde A and Ochayi EO (2020) Leaving No One Behind? Drinking-Water Challenge on the Rise in Niger Delta Region of Nigeria: A Review. Merit Research Journal of Environmental Science and Toxicology (ISSN: 2350-2266) Vol. 6(1): 031–049 DOI: 10.5281/zenodo.3779288

18. Afolabi AS, Raimi MO (2021) When Water Turns Deadly: Investigating Source Identification and Quality of Drinking Water in Piwoyi Community of Federal Capital Territory, Abuja Nigeria. Online Journal of Chemistry, 2021, 1, 38–58; DOI: 10.31586/ojc.2021.010105. www.scipublications.org/journal/index.php/ojc.

19. Morufu OR, Olawale HS, Clinton IE et al., (2021) Quality water not everywhere: Exploratory Analysis of Water Quality Across Ebocha-Obrikom Oil and Gas Flaring Area in the Core Niger Delta Region of Nigeria, 04 October 2021, PREPRINT (Version 1) available at Research Square [https://doi.org/10.21203/rs.3.rs-953146/v1].

20. Raimi MO, Clinton IE, Olawale HS (2021) Problematic Groundwater Contaminants: Impact of Surface and Ground Water Quality on the Environment in Ebocha-Obrikom Oil and Gas Producing Area of Rivers State, Nigeria. Oral Presentation Presented at the United Research Forum. 2^nd^ International E-Conference on Geological and Environmental Sustainability during July 29-30, 2021.

21. Morufu OR, Henry OS, Clinton IE, Gabriel S (2021) Many Oil Wells, One Evil: Potentially toxic metals concentration, seasonal variation and Human Health Risk Assessment in Drinking Water Quality in Ebocha-Obrikom Oil and Gas Area of Rivers State, Nigeria. medRxiv 2021.11.06.21266005; doi: https://doi.org/10.1101/2021.11.06.21266005.

22. Morufu OR, Clinton IE, Bowale A (2021) Statistical and Multivariate Techniques to Trace the Sources of Ground Water Contaminants and Affecting Factors of Groundwater Pollution in an Oil and Gas Producing Wetland in Rivers State, Nigeria. medRxiv 2021.12.26.21268415; doi: https://doi.org/10.1101/2021.12.26.21268415.

23. Raimi MO (2019) 21^st^ Century Emerging Issues in Pollution Control. 6^th^ Global Summit and Expo on Pollution Control May 06-07, 2019 Amsterdam, Netherlands.

24. Raimi MO, Sawyerr HO and Isah HM (2020) Health risk exposure to cypermethrin: A case study of kano state, Nigeria. Journal of Agriculture. 7th International Conference on Public Healthcare and Epidemiology. September 14-15, 2020|Tokyo, Japan.

25. Isah HM, Raimi MO, Sawyerr HO, Odipe OE, Bashir BG, Suleiman H (2020) Qualitative Adverse Health Experience Associated with Pesticides Usage among Farmers from Kura, Kano State, Nigeria. Merit Research Journal of Medicine and Medical Sciences (ISSN: 2354-323X) Vol. 8(8) pp. 432–447, August, 2020. DOI: 10.5281/zenodo.4008682. https://meritresearchjournals.org/mms/content/2020/August/Isah%20et%20al.htm.

26. Olalekan RM, Muhammad IH, Okoronkwo UL, Akopjubaro EH (2020). Assessment of safety practices and farmer’s behaviors adopted when handling pesticides in rural Kano state, Nigeria. Arts & Humanities Open Access Journal. 2020;4(5):191 201. DOI: 10.15406/ahoaj.2020.04.00170.

27. Isah, HM., Sawyerr, HO., Raimi, MO., Bashir, BG., Haladu, S. & Odipe, OE. (2020). Assessment of Commonly Used Pesticides and Frequency of Self-Reported Symptoms on Farmers Health in Kura, Kano State, Nigeria. Journal of Education and Learning Management (JELM), HolyKnight, vol. 1, 31–54. doi.org/10.46410/jelm.2020.1.1.05. https://holyknight.co.uk/journals/jelm-articles/.

28. Morufu OR (2021). “Self-reported Symptoms on Farmers Health and Commonly Used Pesticides Related to Exposure in Kura, Kano State, Nigeria”. Annals of Community Medicine & Public Health. 1(1): 1002. http://www.remedypublications.com/open-access/self-reported-symptoms-on-farmers-health-and-commonly-used-pesticides-related-6595.pdf. http://www.remedypublications.com/annals-of-community-medicine-public-health-home.php.

29. Morufu OR, Tonye VO, Ogah A, Henry AE, Abinotami WE (2021) Articulating the effect of Pesticides Use and Sustainable Development Goals (SDGs): The Science of Improving Lives through Decision Impacts. Research on World Agricultural Economy. Vol 2, No. 1. DOI: http://dx.doi.org/10.36956/rwae.v2i1.347. http://ojs.nassg.org/index.php/rwae/issue/view/31.

30. Hussain MI, Morufu OR, Henry OS (2021) Patterns of Chemical Pesticide Use and Determinants of Self-Reported Symptoms on Farmers Health: A Case Study in Kano State for Kura Local Government Area of Nigeria. Research on World Agricultural Economy. Vol 2, No. 1. DOI: http://dx.doi.org/10.36956/rwae.v2i1.342. http://ojs.nassg.org/index.php/rwae/issue/view/31

31. Hussain MI, Morufu OR, Henry OS (2021) Probabilistic Assessment of Self-Reported Symptoms on Farmers Health: A Case Study in Kano State for Kura Local Government Area of Nigeria. Research on World Agricultural Economy. Vol 2, No. 1. DOI: http://dx.doi.org/10.36956/rwae.v2i1.336. http://ojs.nassg.org/index.php/rwae-cn/article/view/336/pdf.

32. Olalekan RM, Omidiji AO, Williams EA, Christianah MB, Modupe O (2019). The roles of all tiers of government and development partners in environmental conservation of natural resource: a case study in Nigeria. MOJ Ecology & Environmental Sciences 2019;4(3):114□121. DOI: 10.15406/mojes.2019.04.00142.

33. Raimi MO, Tonye VO, Omidiji AO, Oluwaseun EO (2018) Environmental Health and Climate Change in Nigeria. World Congress on Global Warming. Valencia, Spain. December 06-07, 2018.

34. Morufu OR, Tonye VO & Adedoyin OO (2021) Creating the Healthiest Nation: Climate Change and Environmental Health Impacts in Nigeria: A Narrative Review. Scholink Sustainability in Environment. ISSN 2470-637X (Print) ISSN 2470-6388 (Online) Vol. 6, No. 1, 2021 www.scholink.org/ojs/index.php/se. URL: http://dx.doi.org/10.22158/se.v6n1p61. http://www.scholink.org/ojs/index.php/se/article/view/3684.

35. Koleayo OO, Morufu OR, Temitope OW, Oluwaseun EO, Amos LO (2021) Public Health Knowledge and Perception of Microplastics Pollution: Lessons from the Lagos Lagoon, 10 May 2021, PREPRINT (Version 1) available at Research Square [https://doi.org/10.21203/rs.3.rs-506361/v1].

36. Omoyajowo K., Raimi M., Waleola T., Odipe O., Ogunyebi A (2022) Public Awareness, Knowledge, Attitude and Perception on Microplastic Pollution around Lagos Lagoon. Ecological Safety and Balanced use of Resources, 2(24), 35–46. https://doi.org/10.31471/2415-3184-2021-2(24)-35-46.

37. Ikebude, CF (2017). Feasibility Study on Solid Waste Management in Port Harcourt Metropolis: Causes, Effect and Possible Solutions. Nigerian Journal of Technology (NIJOTECH) Vol. 36, No. 1, January 2017, pp. 276 – 281

38. Imam, A., Mohammed, B., Wilson, DC., & Cheeseman, CR. (2008). Solid waste management in Abuja, Nigeria. Waste Management, 28(2), 468–472. https://doi.org/10.1016/j.wasman.2007.01.006.

39. Ogbonna, DN., Amangabara, GT., & Ekere, TO. (2007). Urban solid waste generation in Port Harcourt metropolis and its implications for waste management. Management of Environmental Quality, 18(1), 71–88. https://doi.org/10.1108/14777830710717730.

40. Raimi MO, Adeolu AT, Enabulele CE, Awogbami SO (2018) Assessment of Air Quality Indices and its Health Impacts in Ilorin Metropolis, Kwara State, Nigeria. Science Park Journals of Scientific Research and Impact Vol. 4(4), pp. 060–074, September 2018 ISSN 2315-5396, DOI: 10.14412/SRI2018.074.http://www.scienceparkjournals.org/sri/pdf/2018/September/Olalekan_et_al.pdf. http://www.scienceparkjournals.org/sri/Content/2018/September/2018.htm.

41. Raimi MO, Adio ZO, Odipe OE, Timothy KS, Ajayi BS & Ogunleye TJ (2020) Impact of Sawmill Industry on Ambient Air Quality: A Case Study of Ilorin Metropolis, Kwara State, Nigeria. Energy and Earth Science Vol. 3, No. 1, 2020. URL: http://dx.doi.org/10.22158/ees.v3n1p1. www.scholink.org/ojs/index.php/ees ISSN 2578-1359 (Print) ISSN 2578-1367 (Online)

42. Raimi, OM., Samson, TK., Sunday, AB., Olalekan, AZ., Emmanuel, OO., & Jide, OT. (2021). Air of Uncertainty from Pollution Profiteers: Status of Ambient Air Quality of Sawmill Industry in Ilorin Metropolis, Kwara State, Nigeria. Research Journal of Ecology and Environmental Sciences, 1(1), 17–38. DOI: 10.31586/rjees.2021.010102. Retrieved from https://www.scipublications.com/journal/index.php/rjees/article/view/60.

43. Yusuf OR, Adewoye SO, Sawyerr HO, Morufu OR (2022) Incidence of Hepatitis B and C Viruses among the Scavengers in Kwara State, Nigeria. medRxiv preprint doi: https://doi.org/10.1101/2022.01.26.22269849.

44. Batool S, Nawaz M (2009). Municipal solid waste management in Lahore City District, Pakistan. Waste Manag. 29: 1971-1981. Academia Journal of Environmental Science 4(8): 144–162, August 2016 DOI: 10.15413/ajes.2016.0125

45. Joseph Siji (2015) Waste Collection and Disposal For Optimization of Municipal Solid Lokoja, M.Sc thesis in Partial Fulfilment of the Requirements of the Degree of Master of Science Department of Geography, University of Nigeria, Nsukka.

46. Nachalida Yukalang, Beverley Clarke and Kirstin Ross (2017). Barriers to Effective Municipal Solid Waste Management in a Rapidly Urbanizing Area in Thailand: Int. J. Environ. Res. Public Health 2017, 14, 1013; doi:10.3390/ijerph14091013 www.mdpi.com/journal/ijerph.

47. Kaoje A. U, Sabir AA, Yusuf S., Jimoh AO. and Raji MO (2017). Residents’ perception of solid waste disposal practices in Sokoto, Northwest Nigeria. African Journal of Environmental Science and Technology. Vol. 11(2), pp. 94–102, February 2017 DOI: 10.5897/AJEST2014.1791.

48. Fagariba C, Song S (2016). Assessment of Impediments and Factors Affecting Waste Management: A Case of Accra Metropolis.

49. Steven J (2016). Occupational Risks Associated with Solid Waste Management in the Informal Sector of Gweru, Zimbabwe Journal of Environmental and Public Health, 2016, 9024160 - June 2016. https://doi.org/10.1155/2016/9024160.

